# Clinical Pattern and Outcome of Patients with Acute Kidney Injury in the Emergency Department of Saint Paul’s Hospital Millennium Medical College

**DOI:** 10.1101/2024.02.25.24303349

**Authors:** Berihu Assefa, Yemane Gebremedhin, Benyam Bahta, Frehiwot Worku, Dirijit Mamo, Menbeu Sultan, Mohammed Kalifa

## Abstract

**Background:** Worldwide, 13.3 million people experience Acute Kidney Injury (AKI) each year. 85% of individuals impacted are thought to reside in underdeveloped nations. AKI continues to be one of the most widespread diseases in the world, although little is known about its clinical profile or outcome. The ability to pinpoint particular causes enables the implementation of targeted therapy and the development of preventative measures.

The main goal of this study was to identify the patterns and outcomes of patients with AKI in the emergency department of Saint Paul’s Hospital Millennium Medical College (SPHMMC).

**Method and materials:** A cross-sectional study was conducted at the emergency department of SPHMMC in Addis Ababa, Ethiopia, from June 1-2021 to June 1-2022. Google Forms was used to collect the data, which was then cleaned up in Microsoft Excel before being sent to SPSS version 25 for analysis. To evaluate demographic, clinical profile, and outcome determinants, descriptive statistics, and binary logistic regression analysis were utilized. A paired samples T-test was used to compare the patient’s laboratory findings at admission and discharge.

**Results:** Among the 222 AKI patients included in the study 110 (49.5%) were males and 112 (50.5%) were females. The mean age of presentation was 48±18 years old. The majority of patients were from Addis Ababa (41.4%) and the Oromia region (40.5%). The most common causes of AKI were infections (26.2%), acute glomerulonephritis (20.4%), volume depletion (18.5%), and obstructive uropathy (16.6%). Uremic encephalopathy, infection, malignancy, potassium at discharge from emergency, and low initial Glasgow coma scale (GCS) significantly contributed to the death. The presence of nephrotoxic antibiotics, infection, and hyponatremia significantly contributed to the admission rate to the wards and intensive care unit (ICU).

**Conclusion:** In conclusion, infection is the dominant cause and mortality predictor of AKI at SPHMMC. The majority of patients with infections were sepsis (78.1%), pyelonephritis (11.4%), and pneumonia (10.3%). Early initiation of antibiotics in the emergency is better for a good outcome.

## Background

Acute kidney injury is defined as a decline in renal function over hours or days resulting in the accumulation of toxic wastes and the loss of internal homeostasis. AKI is defined according to the Kidney Disease Improving Global Outcomes (KDIGO) 2012 criteria as an increase in serum creatinine (Scr) by ≥0.3 mg/dl within 48 hours, or an increase in Scr to ≥1.5 times baseline within the prior 7 days, and urine volume < 0.5 mL/ kg/h for 6 hours (1).

The baseline serum creatinine used for hospital-acquired AKI is the first documented serum creatinine on admission. For community-acquired AKI, a Prehospital record of creatinine within seven days to three months of hospital admission, whichever is available, or the minimum and/or most recent value of admission serum creatinine is used as baseline (2).

The acute kidney injury network (AKIN) proposed in 2007, AKI is a decline in kidney function during 48 h as demonstrated by an increase in serum creatinine of more than 0.3 mg/dl, an increase in serum creatinine of more than 50 %, or the development of oliguria (3–7).

AKI affects approximately 13.3 million individuals globally per year. An estimated 85% of those affected live in the developing world. AKI is thought to contribute to about 1.7 million deaths every year (8, 9).

Acute kidney injury (AKI) is a challenging problem in Africa because of the high burden of infectious diseases, the late presentation of patients to healthcare facilities, and the lack of resources to support patients with established AKI (10, 11).

The clinical importance of AKI is exemplified by data showing a consistent association with increased long-term risk of poor outcomes, including death, incident chronic kidney disease (CKD), and greater utilization of health resources. AKI is a common complication of critical illness, with some research showing that as high as 1 in 5 adults experience AKI per hospital admission. The clinical and public health importance of AKI is well established due to its association with high mortality and its separate independent effect on the risk of death and resource use (12).

The International Society of Nephrology conducted a "Global Snapshot" about AKI in 2014, where most cases (45%) were from low and lower-middle-income countries. However, these data may be incomplete, since many of the low-income nations lack the resources to maintain national disease registries, and it is challenging to accurately report cases of AKI outside of major health centers. The incidence of AKI in Low-income countries is not completely understood the proposed reasons being a late presentation of patients to tertiary centers, underreporting, and a reduced capacity to provide intensive care to severely ill patients(13).

## Objectives

General objective

To assess the pattern and outcome of acute kidney injury patients in emergency of SPHMMC from June 1-2021 to June 1-2022.

Specific objectives

- To determine clinical characteristics of AKI in the emergency department of SPHMMC from June 1-2021 to June -2022.
- To explore management practice for AKI patients in the emergency department of SPHMMC, Addis Ababa, Ethiopia from June 1-2021 to June 1-2022.
- To determine the clinical outcome of AKI in SPHMMC emergency from June 1-2021 to June 1-2022.

## Methods and Materials

### Study area

This study was conducted at the emergency of SPHMMC, which is located in Addis Ababa the capital city of Ethiopia. The hospital was established in 1968 by the late Emperor Haile Selassie as Saint Paul’s Hospital. It is a specialized teaching hospital located in the northwestern part of Addis Ababa, the capital of the country. Its catchment population is well over 7 million, making it one of the largest referral centers in the country. As a tertiary center, it receives patients from every corner of the country, most of whom come from the central areas of the country.

In 2010, the hospital was advanced to a medical school, through the decree of the national council of the ministries, and with this, it attained its current name: then got the name, Saint Paul’s Hospital Millennium Medical College (SPHMMC). Being one of the leading tertiary and teaching hospitals in the country, SPHMMC has around 250 faculty members and more than 2800 clinical, academic, administrative & supportive staff. While its inpatient capacity is more than 700 beds, the hospital gives service to an average of 1200 emergency and outpatient clients daily. The hospital serves as a teaching institution for undergraduate and postgraduate medical students and is a research center (14).

The department of emergency and critical care is in charge of its adult emergency. Eight nephrologists work for the nephrology unit, which also offers fellowships. The first and only living-related kidney transplant program was established in September 2015 in collaboration with the University of Michigan (15). The ICU, the kidney transplant department, and the first floor of the maternity block all have areas for dialysis at the hospital.

### Study design

Hospital-based cross-sectional study over one year was conducted by reviewing the charts of patients admitted and diagnosed to have AKI in the emergency department of SPHMMC from June 1-2021 to 1-June 2022.

### Study period & duration

Data collection was carried out from August 1-2022 to September 1-2022 for patients who has been seen at the emergency from June 1-2021 to June 1-2022.

## Population

### Source population

Records of all patients admitted to the emergency department of SPHMMC from Jun 1-2021 to Jun 1-2022.

#### Study population

All patients with acute kidney injury presented to the emergency department of SPHMMC from June 1-2021 to June 1-2022. 222 patients were included in this study.

#### Eligibility criteria

#### Inclusion criteria

Patients who were presented to the emergency department of SPHMMC and diagnosed to have acute kidney injury from Jun 1-2021 to Jun 1-2022 were eligible and included in this study.

#### Exclusion criteria

Those patients with incomplete chart records, established chronic kidney disease (CKD), age under 14 years old, AKI on CKD, multiple admissions during the study period were excluded.

#### Sample size determination and sampling procedure

#### Sample size determination

A simple random sampling technique was employed to select 222 patients from the total of 504 AKI patients who were estimated to be seen over a year. For this study, the medical records of sampled patients that fulfill the inclusion criteria and were presented to the emergency department of SPHMMC from June 1-2021 to June 1-2022 were reviewed.

The following assumptions were considered in calculating the sample size. P value is taken as 0.3 (a mortality rate of AKI patients in a study done in the same city and the same setting) (16). And 95% confidence interval was used, with a 5% margin of error tolerated; the sample size was determined by using single population proportion formula to have the final sample size of 222 patients.

## Study variable

### Dependent variables

#### Outcome

✓ Death
✓ Admitted to wards, ICU, or
✓ Discharged improved

### Independent variables

✓ Age
✓ Sex
✓ Etiology
✓ Precipitating factors
✓ Other co-morbidities (HTN, DM, BPH, pelvic tumors, cardiac disease)
✓ medications

### Operational definition

#### Outcome

conditions of patients written on patients’ chart at discharge time from emergency (admitted to ward or ICU, death, transferred to another hospital, discharged home).

#### Admission

is transferring of a patient from ER to the ward or ICU

#### Disease Pattern

a factor determining and influencing the frequency and distribution of disease, injury, and other health-related events and their cause in a defined human population at admission.

#### Acute kidney injury

Acute kidney injury is defined as a decline in renal function over hours or days resulting in the accumulation of toxic wastes and the loss of internal homeostasis. (17)

#### Adult

refers to individuals above the age group of 14, which is the cut of age for patients to be seen by an emergency department (ED) physician at SPHMMC.

### Data Collection Procedures

Data was collected from all eligible acute kidney injury patients’ charts using a structured checklist by google form. All sampled acute kidney injury patients’ data were collected from the SPHMMC registration logbook, patients’ charts, Health Management Information System (HMIS), referral papers, and death reports. Patient charts and referral papers were reviewed for the age of patients, sex, comorbid illness, medications history, clinical presentation, interventions provided in the hospital, the outcome of patients after the treatment, complications, length of stay, and for other necessary data using a structured checklist in the google form. The checklist was prepared from the articles in the literature review, and guidelines on AKI. The data collection tool was prepared in the English language by and was filled by well-oriented SPHMMC emergency and critical care medicine resident physicians who were not involved in the management of the patients.

### Data quality management

One week before data collection, a 10% pretested was utilized to gather data. However, the checklist needed to be modified to incorporate a few factors. Before starting the data-collecting procedure, the data collectors received training on how to collect data. Close monitoring was kept throughout the data collection process, and the primary investigator and data collectors double-checked full checklists each day to ensure consistency and completeness before analysis.

### Data processing and analysis

After being exported from Google Forms, the data were cleaned up and edited in Microsoft Excel. The IBM SPSS (Statistical Package for Social science) version 25 was then used for data entry, code clearance, and analysis. To look for missing values and variables, frequency and cross-tabulation were utilized. Using descriptive statistics like mean, percentage, frequency, and standard deviation, the demographic and clinical features of the patients were calculated. Paired binary and t-test to identify relationships between independent and dependent variables, logistic regression was performed. Diagrams, tables, and figures are used to present the study’s findings, as seen in the outcome section.

## Results

### Demographic, and clinical characteristics of patients

Of 222 AKI patients identified 112(50.5%) were female patients. The maximum age of the patient identified was 90 years old, and the minimum age identified was 15 years. Most of the patients173 (77.9%) were less than 65 years. Most of patients were from Addis Ababa 92(41.4%) followed by Oromia region 90(40.5%). More than two third (77.5%) of patients were referred from government health facilities. Whereas, 12.6% of patients were referred from private health facilities. The rest 9.9% of patents were self-referral. All the self-referred patients were from Addis Ababa and Oromia region. See Table 1.

**Table 1.** Demographic characteristic of AKI patients presented to the ED of SPHMMC from June 2021 to June 2022.

| Demographic characteristics | Sub-variables | Frequency | Percent |
| --- | --- | --- | --- |
| Age groups | 14-24 | 24 | 10.8 |
|  | 25-34 | 29 | 13.1 |
|  | 35-44 | 44 | 19.8 |
|  | 45-54 | 39 | 17.6 |
|  | 55-64 | 37 | 16.7 |
|  | >64 | 49 | 22.1 |
| Address | Addis Ababa | 92 | 41.4 |
|  | Amhara | 16 | 7.2 |
|  | Oromia | 90 | 40.5 |
|  | SNNPR | 18 | 8.1 |
|  | Others | 6 | 2.7 |
| Patient referred from | Self-referral (home) | 22 | 9.9 |
|  | Government health facility | 172 | 77.5 |
|  | Private health facility | 28 | 12.6 |
| Sex | Male | 110 | 49.5 |
|  | Female | 112 | 50.5 |

**AKI-** Acute Kidney Injury, **ED-** Emergency Department, **SNNPR-** Southern Nations Nationalities and Peoples Region, **SPHMMC-** St. Paul's Hospital Millennium Medical College

From the total 222 AKI patients presented to the adult emergency, the majority 141 (63.5%) were triaged to yellow-green, 60 (27.3%) were triaged to orange and 21 (9.46%) were triaged to red area of the emergency.

Of all the patients presented with normal range blood pressure was 112(50.3%). And 26 patients (11.6%) and 72 patients (32.4%) were in the hypotensive and hypertensive range respectively. Only 12 (5.3%) were presented with hypertensive crises.

More than three-fourths of patients presented with normal temperature, whereas 13.4% had a record of fever on presentation. Most patients had random blood sugar (RBS) of 140-180 (58.9%) and 24.7% of patients with AKI presented with RBS of greater than 180 mg/dl. Only 1.3% of patients had hypoglycemia on presentation to the emergency. See Table 2.

**Table 2.**
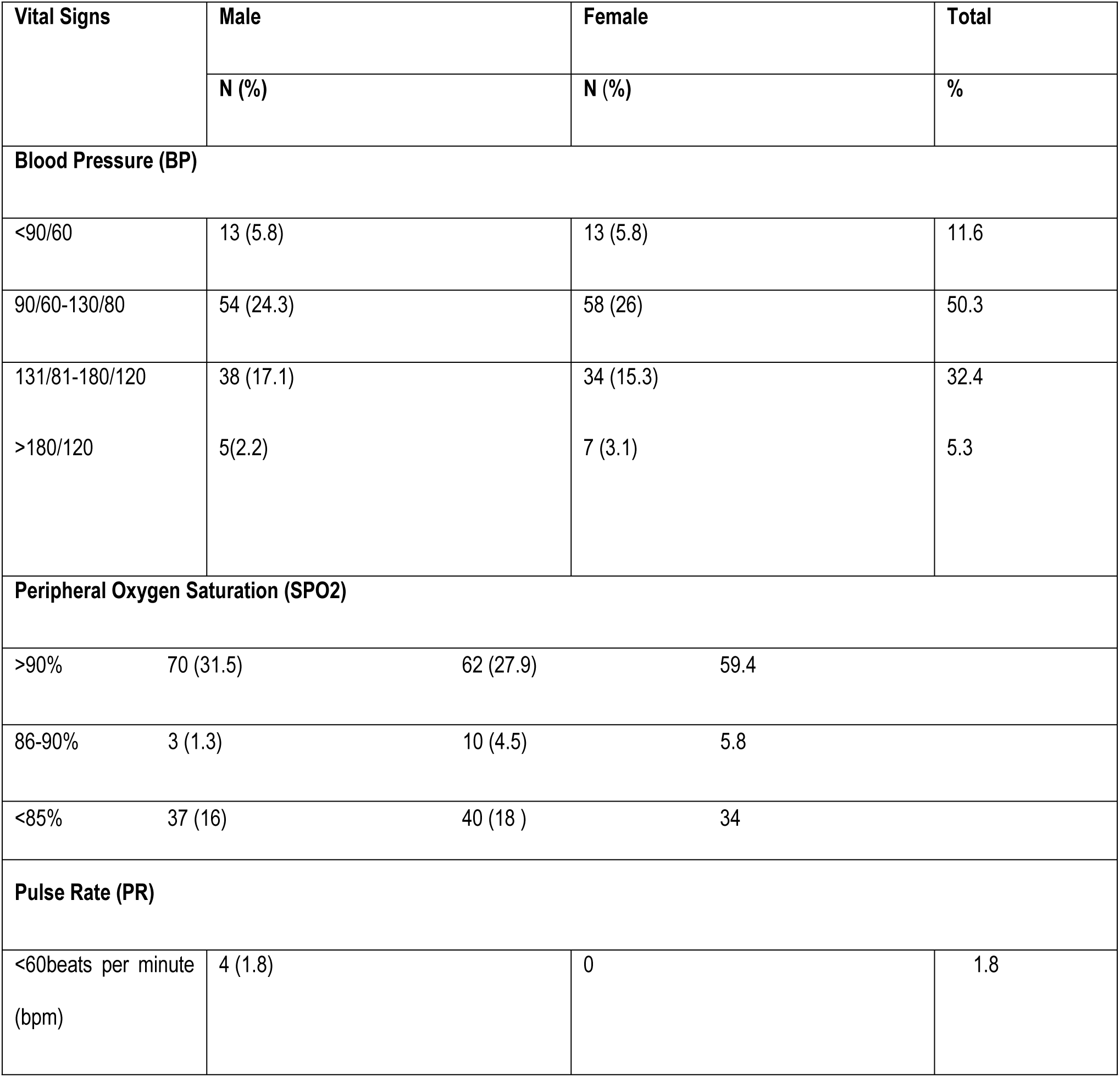
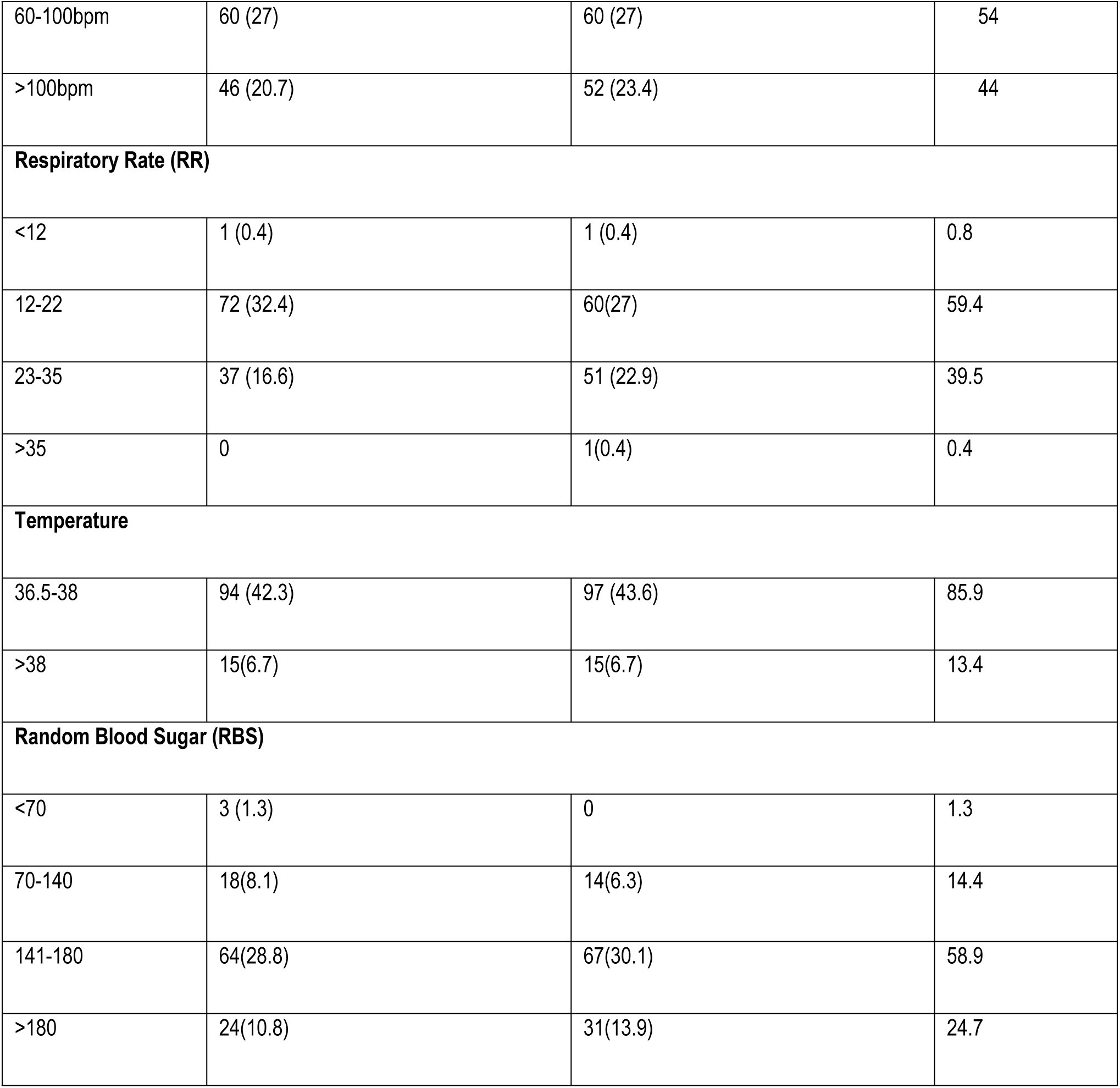
Distribution of vital signs of AKI patients presented to the ED of SPHMMC from June 2021 to June 2022.

### Comorbid conditions

The most common premorbid condition identified was hypertension (27.2 %), followed by malignancy (24.79%) and diabetes mellitus (19%). Heart failure and Human Immunodeficiency Virus (HIV) account for 14% and 8.2% respectively.

### Causes and clinical presentation

The commonest causes of AKI were infection 71 (26.2 %), followed by acute glomerulonephritis 53 (20.4%). Acute glomerulonephritis was not a biopsy, rather a clinical diagnosis with the signs and symptoms and urine analysis as documented by the treating physician. 81% of the patients were fully conscious (GCS=15/15) and 18% had moderately altered mental status (GCS of 9-14). The rest 0.9% of patients presented with coma (GCS=<8) at presentation to the emergency. See Figure 1 below.

**Figure 1:**
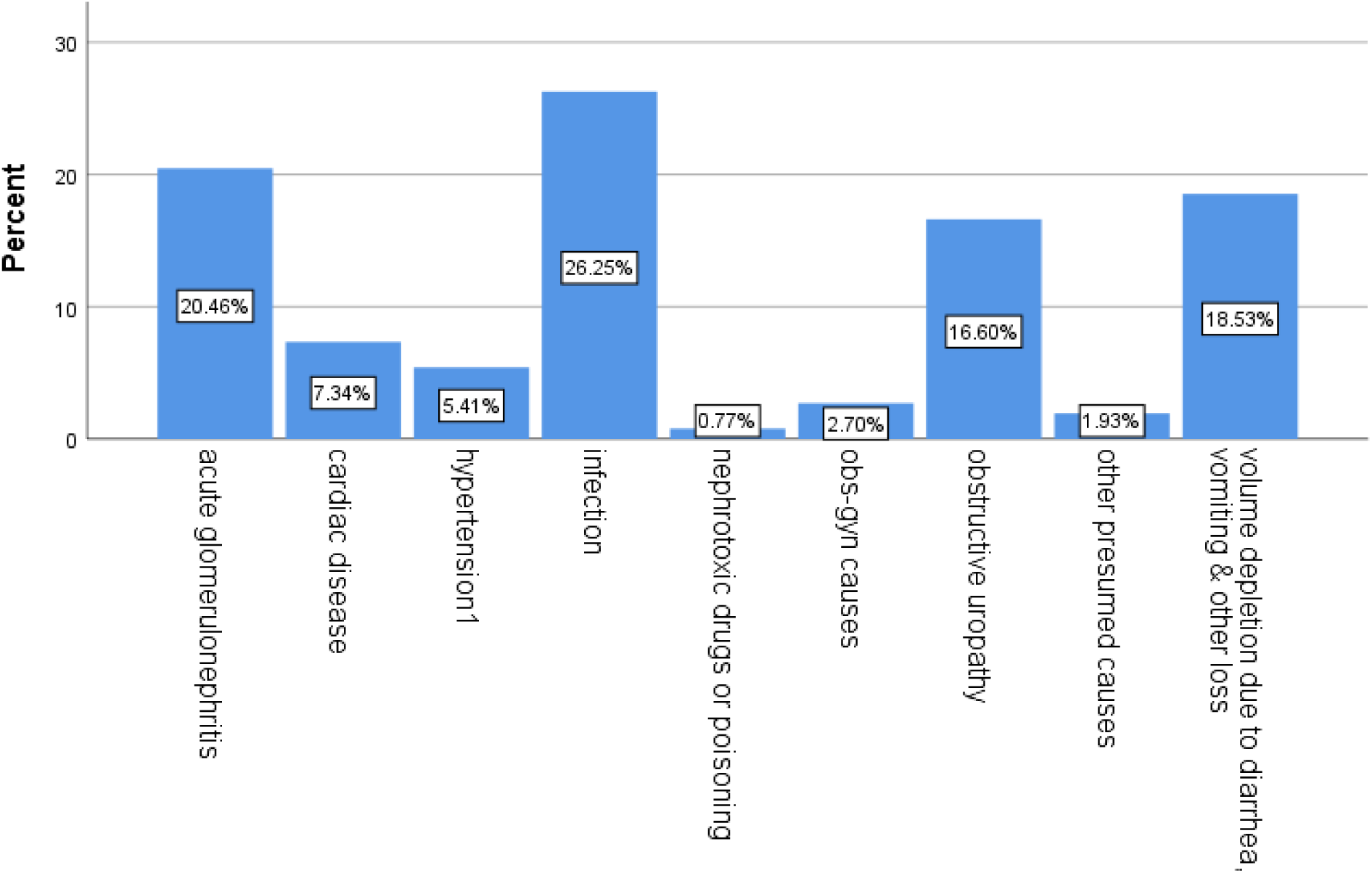
Presumed causes of AKI patients presented to the ED of SPHMMC from June 2021 to June 2022.

The commonest presenting features of AKI were shortness of breath 46 (20.7 %), followed by body edema 44 (19.8 %), decreased urine output 43 (19.3 %), and 13.1 %of AKI patients. See Figure 2 below.

**Figure 2:**
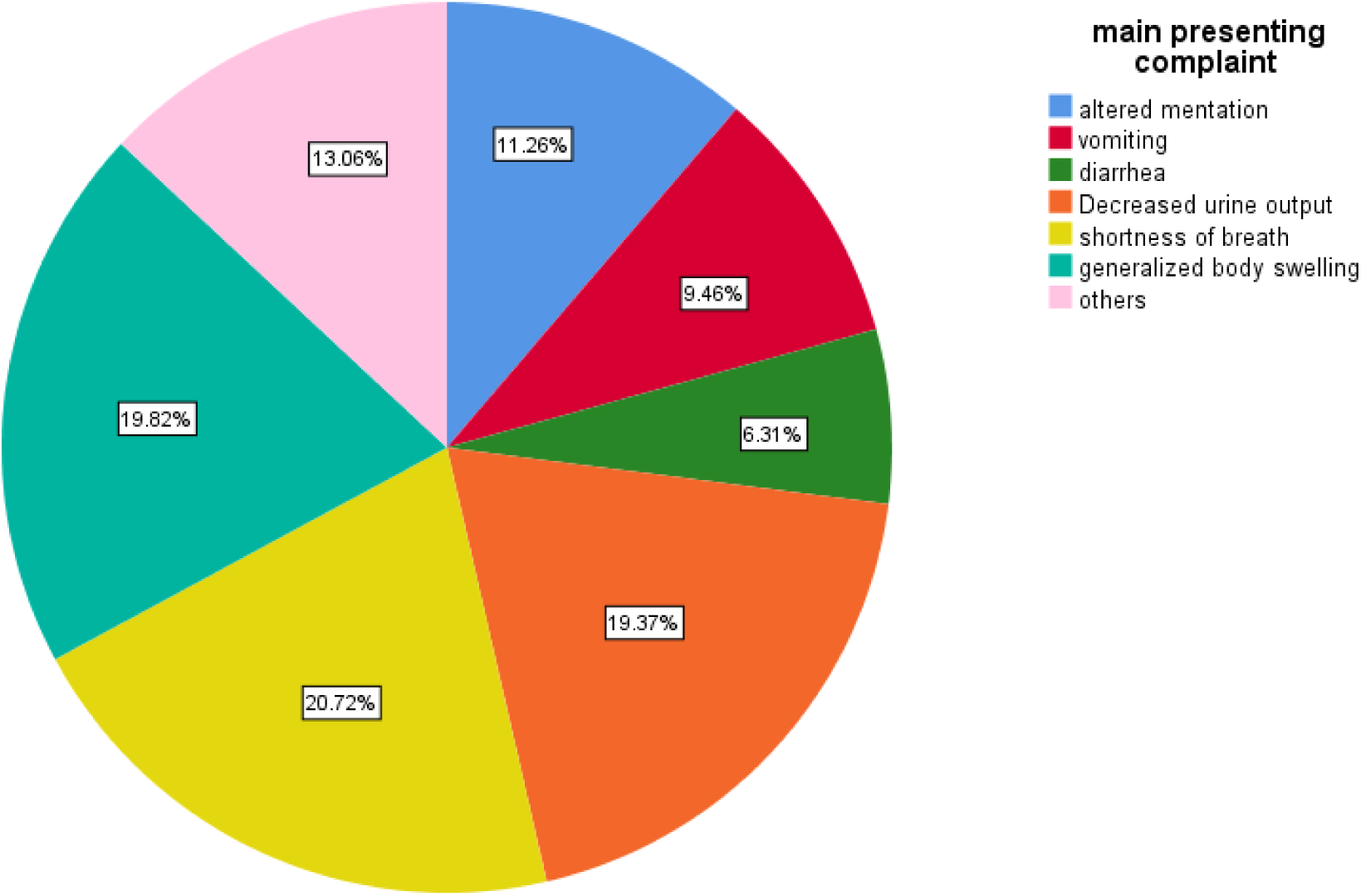
Main presenting complaint of AKI patients presented to the ED of SPHMMC from June 2021 to June 2022.

The most common nephrotoxic drugs traced were antibiotics 28.4% (vancomycin and gentamycin) and immunosuppressive drugs (17.8%) consisting of most anti-cancer drugs like cisplatin. See Table 3 below.

**Table 3:** List of nephrotoxic drugs in AKI patients presented to the ED of SPHMMC from June 2021 to June 2022.

| Nephrotoxic drugs |  |  |  |
| --- | --- | --- | --- |
|  |  | N | Percent |
| Nephrotoxic drugs | Antibiotics | 50 | 28.4% |
|  | NSAIDs | 34 | 19.3% |
|  | Diuretics | 29 | 26.5% |
|  | Immunosuppressive drugs | 49 | 17.8% |
|  | ACEI | 14 | 8.0% |

The most common causes of obstructive uropathy were nephrolithiasis 37.7%, BPH accounting for 35.5%, and malignancy holding 26.6% of all obstructive causes.

Regarding the focus of infection, sepsis accounts for 78.1% of all infection causes, pyelonephritis 11.4%, and pneumonia consist of 10.3%.

Most of the patients (81.3%) had a duration of complaint of two days to two weeks, and 13.6% had more than two weeks duration of complaint.

### Laboratory investigation results

Out of 222 cases, one hundred thirty-seven (61.7%) had normal white blood cell (WBC) on presentation to the emergency, and seventy-five (33.8%) had leukocytosis.

One hundred twenty-six (56.6%) had normal range hemoglobin, and 43.4% of patients were anemic at presentation to the emergency. One hundred-eighty eight (84.5%) patients had elevated initial creatinine, and 137 (61.7%) patients had normal range potassium. Hundred-three (46.4%) patients presented with hyponatremia. See Table 4 below.

**Table 4:** Initial Investigation of AKI patients presented to the ED of SPHMMC from June 2021 to June 2022.

| Variable | Male | Female | Total |
| --- | --- | --- | --- |
| **Hemoglobin Level (Hgb)** |  |  |  |
| Mild anemia | 14(6.3%) | 22(9.9%) | 31(16.2%) |
| Moderate anemia | 21(9.4%) | 28(12.6%) | 49(22%) |
| Severe anemia | 4(1.8%) | 7(3.1%) | 11(4.9 %) |
| Normal | 71(31.9%) | 55(24.7%) | 126(56.6%) |
| **WBC** |  |  |  |
| Leukocytosis | 36(16.2%) | 39(17.5%) | 75(33.7%) |
| Leukopenia | 4(1.8%) | 6(2.7%) | 10(4.5%) |
| Normal range | 70(31.5%) | 67(30.1%) | 137(61.6%) |
| **Platelet** |  |  |  |
| Normal platelet | 48(21.6%) | 58(26.1%) | 120(47.7%) |
| Thrombocytopenia | 55(24.7%) | 47(21.1%) | 102(65.8%) |
| Thrombocytosis | 7(3.1%) | 7(3.1%) | 14(6.2%) |
| **Creatinine** |  |  |  |
| Normal | 17(7.6%) | 17(7.6%) | 34(15.2%) |
| Elevated | 93(41.8%) | 95(42.7%) | 188(84.5%) |
| **Serum Potassium (K+)** |  |  |  |
| Normal | 68(30.6%) | 69(31%) | 137(61.7%) |
| Hypokalemia | 11(4.9%) | 10(4.5%) | 21(9.4%) |
| Hyperkalemia | 31(13.9%) | 33(14.8) | 64(28.7%) |
| **Serum Sodium (Na+)** |  |  |  |
| 135-145 | 53(23.8%) | 51(22.9%) | 104(46.8%) |
| >145 | 7(3.1 %) | 8(3.6%) | 15(6.8%) |
| < 135 | 50(22.5%) | 53(23.8%) | 103 (46.4% |

**Table 5.** Laboratory values of AKI patients presented to the ED of SPHMMC from June 2021 to June 2022.

| Laboratory Results | Mean | Standard Deviation |
| --- | --- | --- |
| Initial WBC | 12219.50 | 10385.291 |
| Discharge WBC | 13689.46 | 21444.763 |
| Initial hemoglobin | 11.87 | 2.880 |
| Discharge hemoglobin | 11.462 | 2.6642 |
| Initial platelet | 263936.94 | 123036.669 |
| Discharge platelet | 273783.33 | 153007.077 |
| Initial blood urea nitrogen ( BUN) | 108.63 | 95.476 |
| Discharge BUN | 92.50 | 84.927 |
| Initial Creatinine | 4.5456 | 4.73147 |
| Discharge Creatinine | 3.608 | 3.8136 |
| Initial K+ | 4.748 | 1.1873 |
| Discharge K+ | 4.458 | .9375 |
| Initial Na+ | 134.50 | 8.953 |
| Discharge Na+ | 136.03 | 7.496 |

The patients had complete blood counts, selected electrolytes, and renal function tests during admission and upon discharge. Most of the patients (84.5%) presented to emergency with elevated creatinine. Only 15.2% of the AKI patients presented with normal creatinine and developed AKI in the hospital.

The mean white cell count is 12219.50 ± 10385.291 and 13689.46 ±21444.763 on admission and discharge respectively. There was good decrement of creatinine upon discharge from a mean of 4.54±4.73 to 3.6±3.81

### Treatment, duration of stay, and complications

More than half (62.1%) received conservative management of drugs, fluids, and urinary catheterization. Around 16% of patients received additional operative management for underlying disease condition. Most of the AKI patients (68.0%) stayed in ED for 1-5 days, 30.1% stayed more than 5 days & 1.8 % of patients stayed less than a day.

Hemodialysis was given for 52 (23.4%) of patients. The identified indications for dialysis were refractory fluid overload (57.6%), uremic complications (30.7%), and refractory hyperkalemia (11.5%).

The most common complication identified in the ED was hyponatremia accounting for 32.5%, hyperkalemia at 22.7%, pulmonary edema at 18.7%, uremic gastropathy at 12.1%, other complications at 7.4 %, and uremic encephalopathy 6.3 % respectively.

Almost half of the patients (50.9%) were admitted to the ward or ICU, one-third (33.3%) were discharged home, twenty-two (9.9%) patients with AKI died in the emergency and (5.9%) were transferred to other hospitals.

The most common immediate cause of death was refractory septic shock (72.7%), hypoxia (22.7%), and massive pulmonary embolism (4.5%).

A paired t-test has been conducted to evaluate the impact of AKI treatment on change in laboratory values during emergency admission and upon discharge from emergency. The result showed a significant decrease in creatinine from 4.5mg/dl (SD 4.7) to 3.6 mg/dl (SD 3.8), t-value of 4.3, and p-0.000. The mean decrement in creatinine was 0.937 mg/dl with a 95% confidence interval ranging from 0.51 to 1.36. There was also a significant change in mean sodium increase from 134 (SD 8.9) to 36 (SD 7.5), t-value of -2.7, and p-value of 0.007. The initial sodium increased with a mean value of 1.5 with a 95% confidence interval ranging from 2.6 to 0.4.

Regarding the WBC count, there was an increase of from 12,219 (SD. 10385) to 13,689 (SD 21,444), t-value of -1.06, and p-value of 0.290 with 95% CI ranging from 4203 to 1263. But it was not a significant change as the p-value is greater than 0.05. On the other hand, there was a significant change in hemoglobin decrement by a mean value of 0.4 mg/dl from the admission value (t-value 3.2, and p-value 0.002 at a 95% CI), ranging from 0.15 to 0.65. There was also a significant change in potassium increment with mean value 0.2896, t-value of 4.2, and p-value of 0.000 with a 95% CI ranging from 0.12 to 0.42. See Table 6 below.

**Table 6.**
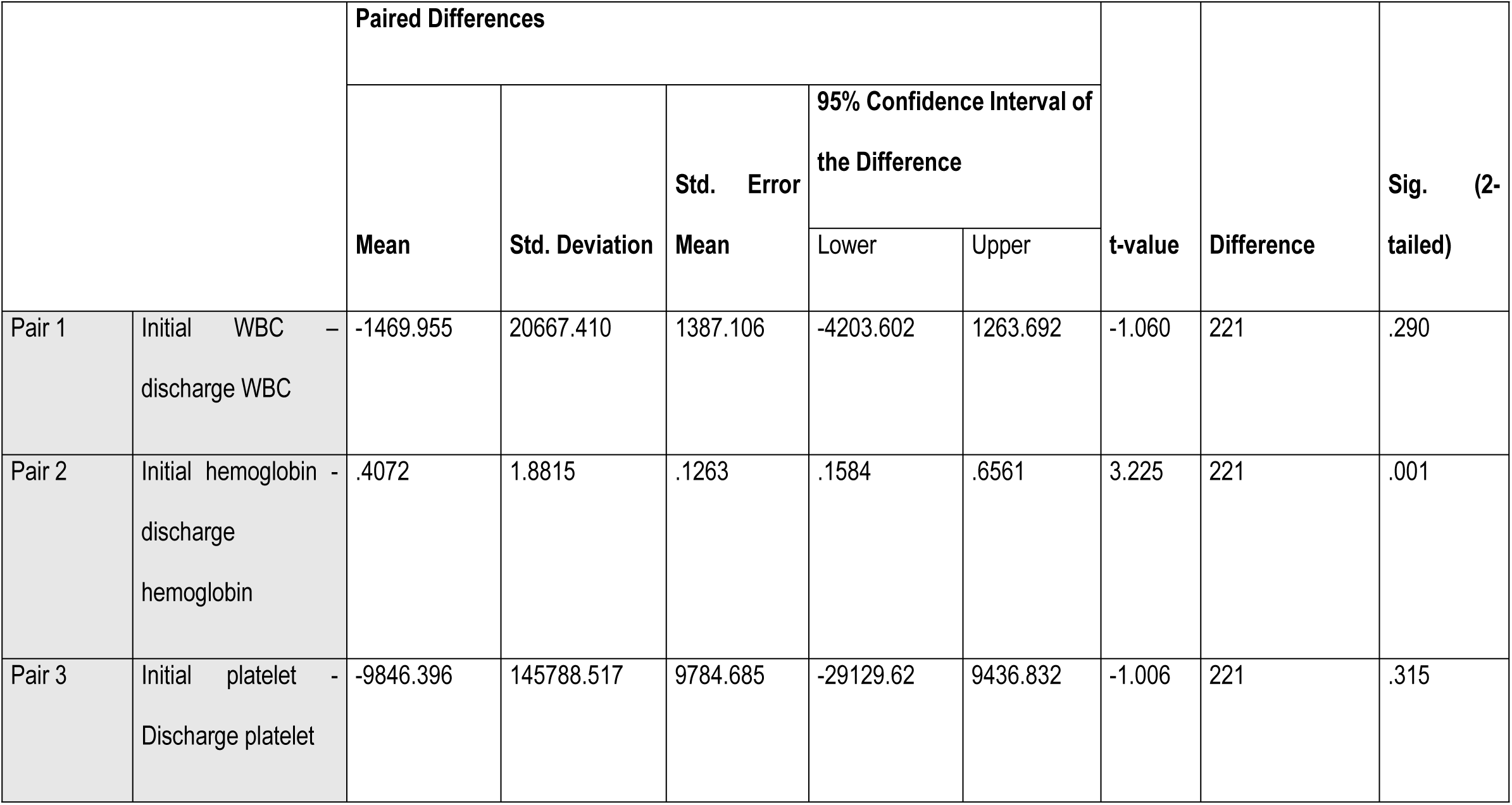
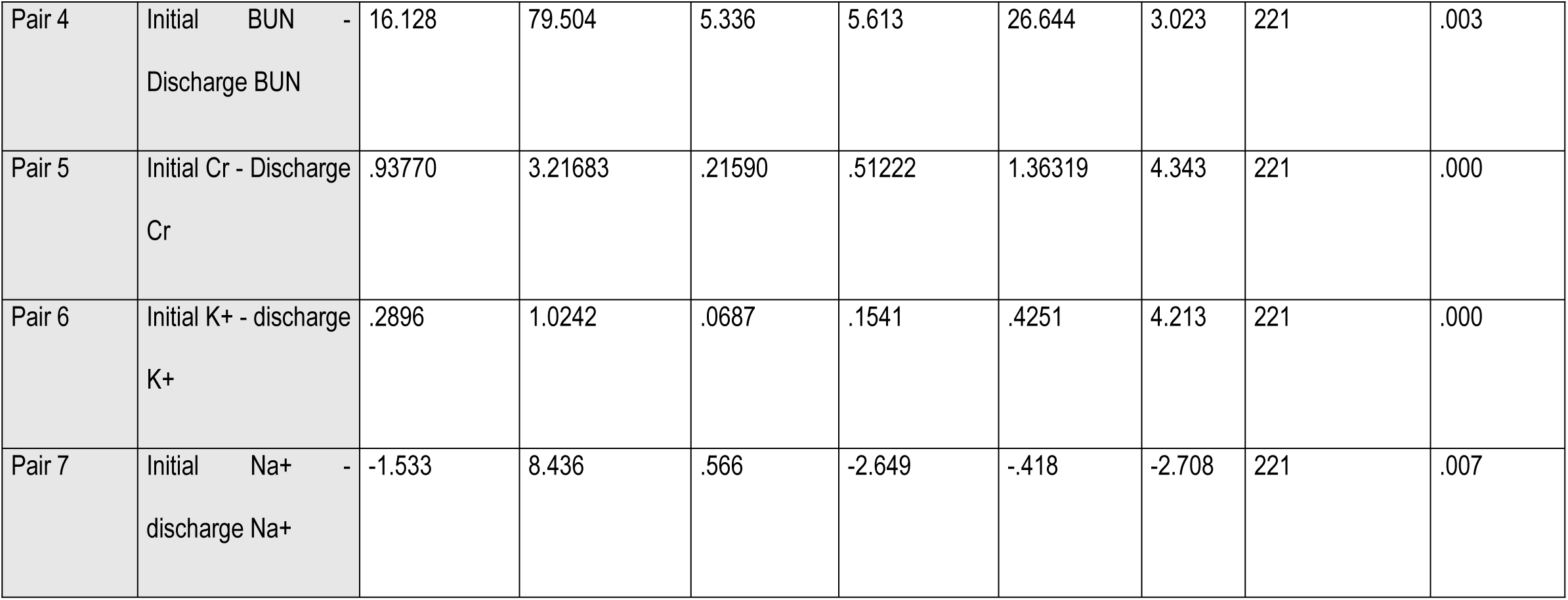
Paired T-test of laboratory values of AKI patients presented to the ED of SPHMMC from June 2021 to June 2022.

### Outcome predictors

Binary logistic regression (95% CI with p-value of < 0.05) was implemented to determine the independent predictors of mortality among AKI patients. Overall AKI patients’ mortality was significantly correlated with the presence of uremic encephalopathy, infection, nephrotoxic-antibiotics, malignancy, and low initial GCS. As shown in Table 7 below.

**Table 7.** Binary logistic regression analysis between different factors and death in AKI patients presented to the ED of SPHMMC from June 2021 to June 2022.

| Death |  |  |  |  |
| --- | --- | --- | --- | --- |
|  | COR (95%CI)) | P-value | AOR (95%CI | P-Value |
| Uremic encephalopathy | 12.9(4.6-36) | 0.000 | 17(5-60) | 0.000 |
| Pulmonary edema | 2.6(1.1-6.5) | 0.03 | **2.1(0.7-6)** | **0.167** |
| Infection | 4(1.7-11.2) | 0.001 | 3.2(1.2-8.5) | 0.017 |
| Discharge K+ | 1.6(1.08-2.5) | 0.002 | **1.54(0.88-2.6)** | **0.126** |
| Nephrotoxic antibiotics | 2.6(1.074-8.7) | 0.035 | 2.7(1.038-7.29) | 0.042 |
| Malignancy | 4.1(1.4-11.9) | 0.007 | 3.5(1.3-9.6) | 0.012 |
| Initial GCS | 4.7(2.06-10.9) | 0.000 | 3.7(1.5-9.06) | 0.003 |

It was also correlated with discharge potassium level and pulmonary edema solely before adjustment of the odds ratio.

Binary logistic regression (95% confidence interval with p-value of < 0.05) was further implemented to determine the independent predictors of admission from emergency to the wards or ICU among AKI patients. In all AKI patients’ admission to the ICU and ward from an emergency was significantly affected by those who were taking nephrotoxic antibiotics, patients with hyponatremia, acute glomerulonephritis, and infection as shown in table 8 below.

**Table 8.** Binary logistic regression analysis between different factors and admission from ED to ward and ICU in AKI patients presented to the ED of SPHMMC from June 2021 to June 2022.

|  | **COR (95%CI))** | **P-value** | **AOR (95%CI))** | **P-value** |
| --- | --- | --- | --- | --- |
| Nephrotoxic antibiotics | 2.5(1.2-4.8) | 0.007 | 0.24(0.11-0.55) | 0.001 |
| Infection | 2.3(1.1-4.3) | 0.014 | 2.1(1.04-4.3) | 0.039 |
| Hyponatremia | 2.1(1.25-3.6) | 0.005 | 0.45(0.25-0.83) | 0.011 |
| Acute glomerulonephritis | 0.39(0.20-0.75) | 0.005 | 0.45(0.22-0.91) | 0.027 |

## Discussion

In the current study, we conducted a cross-sectional study on the clinical profile and outcome of patients with acute kidney injury in adult emergency of SPHMMC from June1-2021 to June-1-2022. The most common presenting complaints are shortness of breath (20.7%), generalized body swelling (19.8%), and decreased urine output (19.37%). the most common identified comorbidities are hypertension (27.2%) and malignancy (24.7%). Infection (26.2%), acute glomerulonephritis (20.46%), volume depletion due to diarrhea & other loss are the most common presumed causes of AKI. In our study, infection consists of sepsis (78.1%), acute pyelonephritis (11.4%) pneumonia (10.3%). Hyponatremia, hyperkalemia, and pulmonary edema are the most common identified complications of AKI identified in the emergency. Nearly half of the patients (50.9%) were admitted to the wards or ICU from the emergency for further management. Of 222 AKI patients, 22 patients passed away in an emergency with the most common cause of death, septic shock.

Most of the published data estimating the incidence of AKI were from high-income countries and included patients admitted to critical care units; settings where the ability to trace and detect suspected cases remains high. In a systematic review, the world incidence of AKI among hospitalized adults was estimated to be 21.6%, and the condition was found to be associated with a 23.9% increase in mortality (18)

This study elucidated the referral area, presenting vital signs, triage categories, main presenting complaint, causes, complications, the practice of management, and outcomes of patients with AKI. The presentation of the patients at a younger age coincided with results from other African countries as well as those done in Ethiopia (10, 19, 20)

Most of our AKI patients are from Addis Ababa and Oromia regions. Most of them were referred from government health facilities, though there are a few patients referred from private health facilities and self-referred. The gender proportion of patients is almost 1:1 with a median age of 47 years But, in the previous study of the same hospital, the mean age was 37 years (16, 19, 21).

Forty-nine patients (22.1%) of the study population were over the age of sixty-four in our study, as opposed to the study conducted by JIMMA Hospital, which included 28% of AKI patients in this age range. AKI-affected patients who were younger than 65 years old made up more than three-fourths of the total patient population (22).

The most frequent comorbid conditions in our study were hypertension (27.2%), malignancies (24%) diabetes mellitus (19%), and heart failure (14%). The comorbidities were hypertension (30.5%), HIV (32.6%), and diabetes (13.3%), which are similar to research conducted in South Africa in 2017. However, research conducted in Uganda found that HIV accounts for 60% of all comorbid conditions (12). This may be due to lower triggering factors to screen HIV in AKI patients. But, HIV, its complications, and the medications taken could cause significant renal derangement.

The most presenting complaint of AKI patients to the emergency was shortness of breath (20.7%), generalized body swelling (19.8%), and decreased urine output (19.3%). Our finding was consistent with a study done in Tanzania emergency (21).

The most common cause of AKI identified in our study were infections, acute glomerulonephritis, and hypovolemia 26%, 20%, and 18.5% respectively. In a previous study done in the same hospital, the commonest cause identified was hypovolemia. Obstructive uropathy accounts for the fourth most common cause of AKI in our study, which is similar to studies done in the same city and same setup. But the most common cause of obstructive uropathy identified in our patients is nephrolithiasis rather than cervical cancer in the comparative studies done in the same study (10, 19, 22).

Regarding AKI-associated complications, hyponatremia (32.5%), hyperkalemia (22.7%), and pulmonary edema (18.7%) are the most common. Though our findings with hyperkalemia and pulmonary edema are similar in other studies (21). Hyponatremia was a different finding in our study.

Of the all patient enrolled in the study 21% of patients required dialysis, and 62% managed conservatively with fluids, antibiotics, steroids, and other methods of management for the cause of AKI. The rest 16.6% of patients were operated on as part of treatment for the underlying cause of AKI. Our finding regarding management is more or less similar to previous studies done in another hospital of the same city and a study done in Tanzania, on an emergency set (16, 21).

The most common indication for dialysis was refractory fluid overload (57.6%), other uremic complications (uremic encephalopathy, uremic pericarditis, uremic gastropathy, and uremic bleeding all accounting for 30.7 % indication for dialysis) and hyperkalemia (11.5%). As compared to other studies done in a similar city and the same setup, uremic encephalopathy (72%) is the most common indication for dialysis, followed by hyperkalemia (27%) and fluid overload (22%). Metabolic acidosis could not be diagnosed in our hospital, as there is no arterial blood gas analysis (16).

Regarding treatment outcome, half (50.9%) of the AKI patients were admitted to the wards or ICU, whereas, one-third of patients were discharged improved and death outcome accounted for 9.9 %. Around 84% of patients had elevated initial creatinine on admission to the emergency and 77% of patients were disposed of with persistently elevated creatinine. Only 23% of patients disposed of resolved AKI from emergency. This indicates that our result is similar to a study done in Ghana where 70-90% of AKI is community-acquired (20). The mortality of patients in the study done in Tanzania, emergency set up was 39 % which is high than our result (21). The same higher result mortality of AKI has been observed study done in TASH which is a tertiary teaching hospital, in the same capital city. It may be due to the inclusion of AKI in CKD patients (16). The mortality rate of 9.9% in our study is much lower than the data from a global meta-analysis of studies done across the globe which has shown a pooled AKI-associated mortality rate of 49.4 % for dialysis requiring AKI (23). But, the result of our death rate closes the range put in the metanalysis done in Ghana, which puts the AKI death rate at 11.5-43.5% (20).

In our current study uremic encephalopathy, infection, malignancy, potassium at discharge from emergency and low initial GCS were significantly contributed to death rate. In the other study done in JIMMA, the duration of AKI, the severity of AKI, and hyperkalemia were associated with the death rate (22).

Similarly, a study done in Sudan showed that increased age, the severity of AKI, presence of sepsis and liver disease all contributed to increased mortality in AKI patients (18).

In the same manner, binary logistic regression showed that the presence of nephrotoxic antibiotics, infection, and hyponatremia significantly contributed to admission rate to the wards and ICU.

## Conclusion and Recommendation

Most developing countries showed similarities in the AKI pattern we analyzed. AKI was primarily a community-acquired condition in most of our patients. Infections, acute glomerulonephritis, and hypovolemia are the most frequent causes of AKI at Saint Paul’s Hospital millennium medical college. Pneumonia (10.3%), pyelonephritis (11.4%), and sepsis (78.1%) were the three most prevalent conditions among the patients, with a fatality rate of 9.9%. Over 50% of patients with AKI were admitted to the wards or ICU. Significant mortality predictors included uremic encephalopathy, infection, malignancy, and low GCS at presentation.

So health care providers should focus on the prevention strategies for the most common contributors of AKI development, mortality, complications, and causes of admission to decrease the health institutions burden of care.

## Strength and limitation

### Strengths

The study assessed clinical patterns and outcomes of AKI patients in adult emergency and we tried to formulate an association between outcomes and predictors affecting outcomes by regression analysis.

### Limitations

This is a single-center cross-sectional observational hospital-based study and the results cannot be generalized to the general population.

## Declarations

### Ethical consideration

The St. Paul’s Hospital Millennium Medical College Institutional Review Board in Addis Ababa provided ethical clearance and permission for this study with Ref. No. pm23/11. The Department of EMCC & Research Directorate sent a letter of authorization to the medical record unit requesting access to and examination of the patient charts. No names or other personal information was utilized in the data collection to ensure confidentiality. The need for informed consent and publication was waived by SPHMMC IRB due to the retrospective nature of data collection.

### Conflict of interest

None.

### Source of funding

Funding was not received for this research, but logistic expenses were covered by Saint Paul’s Hospital Millennium Medical College.

### Data availability

Data were presented without restriction and available upon request to the corresponding author.

### Consent to publish

Not applicable.

## Acknowledgments

I would want to express my profound gratitude to SPHMMC’s department of emergency and critical care medicine for providing me with the opportunity to carry out my research. I would like to express my profound gratitude for my advisors for their insightful output throughout this research.

Last but not least, I want to express my thanks to my parents and my beloved wife for their unwavering support and bravery.

## Notes

### Competing Interest Statement

The authors have declared no competing interest.

### Funding Statement

The author(s) received no specific funding for this work.

### Author Declarations

The St. Paul's Hospital Millennium Medical College Institutional Review Board in Addis Ababa provided ethical clearance and permission for this study with Ref. No. pm23/11. The Department of EMCC & Research Directorate sent a letter of authorization to the medical record unit requesting access to and examination of the patient charts. No names or other personal information was utilized in the data collection to ensure confidentiality.

## References

1. Khwaja A. KDIGO clinical practice guidelines for acute kidney injury. Nephron Clinical Practice. 2012;120(4):c179–c84.

2. KDIGO clinical practice guideline for the care of kidney transplant recipients. American journal of transplantation: official journal of the American Society of Transplantation and the American Society of Transplant Surgeons. 2009;9 Suppl 3:S1–155.

3. Mehta RL, Kellum JA, Shah SV, Molitoris BA, Ronco C, Warnock DG, et al. Acute Kidney Injury Network: report of an initiative to improve outcomes in acute kidney injury. Critical care (London, England). 2007;11(2): R31.

4. Judith. tintinallis emergency medicine2018. 2159 p.

5. Ja M, Hockberger R, Walls R. Rosen’s emergency medicine: concepts and clinical practice2010.

6. Barry R, James MT. Guidelines for classification of acute kidney diseases and disorders. Nephron. 2015;131(4):221–6.

7. Makris K, Spanou L. Acute kidney injury: definition, pathophysiology and clinical phenotypes. The clinical biochemist reviews. 2016;37(2):85.

8. Li PK, Burdmann EA, Mehta RL. Acute kidney injury: global health alert. Kidney international. 2013;83(3):372–6.

9. Mesic E, Aleckovic-Halilovic M, Pjanic M, Hodzic E, Dugonjic-Taletovic M, Halilcevic A, et al. Recent Pattern of Acute Kidney Injury in Bosnia and Herzegovina. Medical Archives. 2019;73(4):276.

10. Caballo B, Khogali M, Khalifa E, KhaIiI E, EI-Hassan A, Abu-Aisha H. Patterns of "Severe Acute Renal Failure" in a referral center in Sudan: Excluding intensive care and major surgery patients. Saudi Journal of Kidney Diseases and Transplantation. 2007;18(2):220–5.

11. Cerdá J, Lameire N, Eggers P, Pannu N, Uchino S, Wang H, et al. Epidemiology of acute kidney injury. Clinical journal of the American Society of Nephrology. 2008;3(3):881–6.

12. Bagasha P, Nakwagala F, Kwizera A, Ssekasanvu E, Kalyesubula R. Acute kidney injury among adult patients with sepsis in a low-income country: clinical patterns and short-term outcomes. BMC Nephrology. 2015;16(1):4.

13. Feehally J. The ISN 0by25 Global Snapshot Study. Annals of nutrition & metabolism. 2016;68 Suppl 2:29–31.

14. Saint paul.s hospital millennium medical college. historical background of hospital and college: SPHMMC; 2022 [Available from: https://sphmmc.edu.et.

15. Ahmed MM, Tedla FM, Leichtman AB, Punch JD. Organ transplantation in Ethiopia. Transplantation. 2019;103(3):449–51.

16. Kefyalew M, Azazh A, Siraw BB, Teklu A, Seid A, Admasu TN. Clinical Profile and Outcome of Acute Kidney Injury: A Prospective Cross-sectional Study at a Tertiary Hospital in Addis Ababa, Ethiopia. 2021.

17. Abebe A, Kebede B, Woobie Y. Clinical Profile and Short-Term Outcomes of Acute Kidney Injury in Patients Admitted to a Teaching Hospital in Ethiopia: A Prospective Study. Int J Nephrol Renovasc Dis. 2021;14:201–9.

18. Yousif DE, Topping AR, Osman MF, Raimann JG, Osman EM, Kotanko P, et al. Acute Kidney Injury in Sub-Sahara Africa: A Single-Center Experience from Khartoum, Sudan. Blood Purification. 2018;45(1-3):201–7.

19. Ibrahim A, Ahmed MM, Kedir S, Bekele D. Clinical profile and outcome of patients with acute kidney injury requiring dialysis—an experience from a hemodialysis unit in a developing country. BMC Nephrology. 2016;17(1):1–5.

20. Riley S, Diro E, Batchelor P, Abebe A, Amsalu A, Tadesse Y, et al. Renal impairment among acute hospital admissions in a rural Ethiopian hospital. Nephrology (Carlton, Vic). 2013;18(2):92–6.

21. Sylvanus E, Sawe HR, Muhanuzi B, Mulesi E, Mfinanga JA, Weber EJ, et al. Profile and outcome of patients with emergency complications of renal failure presenting to an urban emergency department of a tertiary hospital in Tanzania. BMC Emergency Medicine. 2019;19(1):11.

22. Belay M, Kebede B, Woobie Y. Mortality and predictors of acute kidney injury in adults: a hospital-based prospective observational study. 2021:91.

23. Susantitaphong P, Cruz DN, Cerda J, Abulfaraj M, Alqahtani F, Koulouridis I, et al. World Incidence of AKI: A Meta-Analysis. Clinical Journal of the American Society of Nephrology. 2013;8(9):1482–93.

